# Maternal childhood trauma and perinatal distress predict the development of attention in infants from 6 to 18 months in a Swedish cohort study

**DOI:** 10.1101/2021.09.13.21263510

**Authors:** Hsing-Fen Tu, Alkistis Skalkidou, Marcus Lindskog, Gustaf Gredebäck

**Affiliations:** Department of Neurology, Max Planck Institute for Human Cognitive and Brain Sciences, Leipzig, Germany; Department of Women’s and Children’s Health, Uppsala University, Uppsala, Sweden; Department of Psychology, Uppsala University, Uppsala, Sweden

**Keywords:** Sustained attention, infancy, maternal anxiety, maternal childhood trauma, cross-generational effect

## Abstract

Maternal distress is repeatedly reported to have negative impacts on the cognitive development in children. Studies examining the association between maternal distress and the development of attention in infancy are few. This study investigated the longitudinal relationships between maternal distress (depressive symptoms, anxiety symptoms, and exposure to childhood trauma) and the development of attention in infancy in 118 mother-infant dyads. We found that maternal exposure to non-interpersonal traumatic events in childhood and a large degree of anxiety during the 2^nd^ trimester was associated with less attention of the infants to audio-visual stimuli at 6, 10, and 18 months. In addition, exposure to interpersonal traumatic events in childhood was identified as a moderator of the negative effect of maternal anxiety during the 2^nd^ trimester on the development of attention in infants. We discuss the possible mechanisms accounting for these cross-generational effects. Our findings underscore the importance of maternal mental health to the development of attention in infancy and address the need for early screening of maternal mental health during pregnancy.

## Introduction

Attention in infancy is an important cognitive operation that involves alerting, orienting, and attending to information in the environment (Colombo, 2001; Petersen & Posner, 2012). Attention develops rapidly in the first postnatal year (Xie, Mallin, & Richards, 2019) and continues to develop into adulthood (Hoyer, Elshafei, Hemmerlin, Bouet, & Bidet-Caulet, 2021), playing a fundamental role in learning (Holland & Maddux, 2010; Johnson, Posner, & Rothbart, 1991). Attention has also been linked to the development of self-regulation and executive function in childhood, and later in life (Cuevas & Bell, 2014; Posner & Rothbart, 2009). Attention has also been reported to be a predictor of social development (Bowers et al., 2019), cognitive functioning (Lawson & Ruff, 2004), language development (Yu, Suanda, & Smith, 2019), and academic skills (Shannon, Scerif, & Raver, 2021). Poor attention skills are related to several neurodevelopmental disorders, such as attention-deficit/hyperactivity disorder (Barkley, 1997), autism spectrum disorder (Matson, Rieske, & Williams, 2013), and fragile X syndrome (Scerif, Longhi, Cole, Karmiloff-Smith, & Cornish, 2012).

Empirical studies of infants and children have focused on identifying early risk factors that hinder the development of attention. Generally speaking, the development of attention has been suggested to result from the interaction between biological factors and early parental environment (Faraone & Larsson, 2019; Scerif, 2010; Voelker, Sheese, Rothbart, & Posner, 2009). From this perspective, the fetal phase is critical because genetic factors and the in-utero environment are intertwined during early development. Later in the first postnatal year, mothers continue to be the major provider of environmental stimulation in many cultures. Both biological and environmental risk factors are associated with maternal distress (Fawcett, Fairbrother, Cox, White, & Fawcett, 2019; Priest, Austin, Barnett, & Buist, 2008). Operationally, maternal distress is often symptomized by an unbalanced and/or strained emotional state from pregnancy to postpartum and commonly includes depression and/or anxiety (Fontein-Kuipers, 2016; Priest et al., 2008). Moreover, the severity of psychological distress is strongly linked to exposure to traumatic experiences earlier in life, such as natural disaster, physical harm, and violence, among others (Chu, Williams, Harris, Bryant, & Gatt, 2013; Sexton, Hamilton, McGinnis, Rosenblum, & Muzik, 2015). Some studies have suggested that different types of traumatic events, such as interpersonal and non-interpersonal trauma, have different impacts on mental distress and psychiatric symptoms (Baker et al., 2020; Haldane & Nickerson, 2016).

Evidence supporting an association between maternal distress and neurological development indicates changes in cortical and subcortical connectivity in human infants (Rifkin-Graboi et al., 2013; Scheinost, Spann, McDonough, Peterson, & Monk, 2020) and a negative impact on neurogenesis and gene expression in neonates of rodents (Fatima, Srivastav, Ahmad, & Mondal, 2019). Negative impacts on cognitive development in human children have also been demonstrated in several studies. For example, Laplante et al. (2004) reported that prenatal maternal distress in the 1^st^ trimester of pregnancy is associated with lower general intellectual and language development at the age of 2 years. Keim et al. (2011) demonstrated that increasing anxiety levels from pregnancy to postpartum is linked to a decrease in cognitive function at the age of 12 months. Furthermore, a meta-analysis of 11 studies reported a small effect size for the association between maternal distress in the 3^rd^ trimester and early child cognitive development (Tarabulsy et al., 2014). In a review of toddlers’ cognitive development, 6 out of 7 moderate- to strong-quality studies suggested a negative association with prenatal maternal distress; 4 out of 5 studies demonstrated similar results for postnatal maternal distress. In the same review, one study including prenatal and postnatal maternal distress suggested that the two time points independently affect toddlers’ cognitive development (Kingston, McDonald, Austin, & Tough, 2015; Koutra et al., 2013).

To date, evidence from empirical studies has also indicated that maternal distress might have negative impact on attention-related development in infants of both human and nonhuman primates. In nonhuman primates, exposure to mild stress during pregnancy is related to less visual exploration and higher distractibility of offspring (Schneider, 1992). In humans, maternal stress during pregnancy has been reported to have a negative impact on infants’ attention shifting at the age of 18 months (Plamondon et al., 2015). A similar negative impact from stress during pregnancy was reported by Merced-Nieves et al. (2020), who showed that infants whose mothers perceived higher stress during pregnancy needed more time than others to process visual information at the age of 7.5 months and looked away from the tasks significantly more than infants whose mothers had low perceived stress during pregnancy (Merced-Nieves, Dzwilewski, Aguiar, Lin, & Schantz, 2020). In addition, preliminary evidence suggests that this cross-generational association between maternal distress and infant development is possibly linked or mediated by trauma exposure prior to pregnancy (Bosquet Enlow et al., 2017; Bouvette-Turcot et al., 2020).

Maternal distress during the postnatal phase also affects infants’ attention. For example, infants of depressed mothers have been observed to have less synchronous mutual gaze with their mothers than infants of non-depressed mothers (Lotzin et al., 2015). In turn, mutual gaze has been associated with visual attention in the first postnatal year of life (Niedźwiecka, Ramotowska, & Tomalski, 2018). The impact of maternal distress on mother-infant interactions (Granat, Gadassi, Gilboa-Schechtman, & Feldman, 2017) and maternal sensitivity (Bernard, Nissim, Vaccaro, Harris, & Lindhiem, 2018) have been related to infants’ selective attention (Juvrud, Haas, Fox, & Gredebäck, in press) and gaze-following ability (Astor et al., 2020).

Maternal distress also influences the developmental trajectories of attention in childhood. Several studies including large cohort groups have reported that maternal depressive and anxiety symptoms are significantly associated with attention problems in offspring at the ages of 2 years (Ross, Letourneau, Climie, Giesbrecht, & Dewey, 2020), 3 and 4 years (Van Batenburg-Eddes et al., 2013), as well as 5, 6.5, and 14 years (Clavarino et al., 2010; Wang & Dix, 2017). Two large cohort studies measuring the associations between maternal distress and ADHD symptoms at the age of 4 and 8–9 years, respectively, demonstrated similar results (Mulraney et al., 2019; Vizzini et al., 2019). The link between maternal distress and attention problems in offspring continues into adolescence (Ayano, Betts, Tait, Dachew, & Alati, 2021).

Regarding maternal lifetime trauma, two large cohort studies reported that maternal childhood abuse and adverse experiences contribute to ADHD in offspring (Moon, Bong, Kim, & Kang, 2021; Roberts, Liew, Lyall, Ascherio, & Weisskopf, 2018). Roberts et al. (2013) also observed an association between maternal childhood abuse exposure and an elevated risk of autism in their children.

Overall, the literature suggests that maternal mental health contributes to the development of attention in children. However, contrary to systematic and compelling evidence indicating the impact of specific variables related to maternal psychological distress and maternal lifetime trauma exposure on the development of attention in childhood, there is only very little evidence in infancy. Moreover, most previous studies examined the impact of one or two aspects of maternal distress on infants or children’s attention, we took depression, anxiety, and trauma exposure into consideration. The analysis of multiple risk factors together is essential due to the high likelihood of co-morbidity and high correlations between risk factors. To bridge this gap, the main aim of this longitudinal study was to investigate how maternal childhood trauma and maternal distress (depression and anxiety from antenatal 17, 32 weeks to postpartum 6 weeks and 6 months) are related to infants’ attention from 6 to 18 months of age. We hypothesize that maternal childhood trauma exposure and maternal distress are negatively associated with the infant’s attention. From our understanding, this is the first study to assess the full path from trauma prior to pregnancy to maternal distress during pregnancy to infancy, and its effect on infants’ attention.

If an association between maternal distress and infants’ attention was confirmed, we planned to further explore potential buffers. As the literature addressing this question is scarce, we narrowed our focus to three dimensions of protective factors for which there is some literature in humans suggesting that a buffering effect is present, even if not directly demonstrated in prior work. The first dimension is the subjective delivery expectations and experience of mothers, which is associated with different trajectories of perinatal depression and the temporal onset of postnatal depression (Wikman et al., 2020). The second dimension is the amount of sleep that mothers get during pregnancy and postpartum, which has previously been shown to be associated with maternal distress (Palagini et al., 2014; Tikotzky, 2016). The third dimension focused on the interpersonal perspective, including the partner’s social support and mother-infant bonding. Regarding the partner’s support, supportive partnerships, e.g., social support and pregnancy-specific received support, has been observed to be linked to both reduced maternal postpartum distress and infants’ distress to novelty (Stapleton et al., 2012). Reduced maternal distress is also linked to positive mother-infant bonding [e.g., breast feeding (Adedinsewo et al., 2014) or bounding difficulties (Dubber, Reck, Müller, & Gawlik, 2015; Fransson et al., 2020)]. In preterm infants, daily skin-to-skin contact between the newborn and mother has been reported to reduce the paternal stress in the first postnatal year and was linked to better cognitive control of their children at the age of 10 years (Feldman, Rosenthal, & Eidelman, 2014). Taken together, we expect that positive delivery expectations and experiences, sufficient maternal sleep, and positive mother-infant bonding are positive factors that can buffer against the negative effect of maternal distress on infants’ attention.

## Methods

### Participants

The final data included 118 mother-infant dyads from the BASICchild cohort as part of a longitudinal study (the Basic Child project; Gredebäck, Forssman, Lindskog, & Kenward, 2019) of a subsample of the population-based BASIC study (“Biology, Affect, Stress, Imaging, and Cognition (BASIC)”, Axfors et al., 2019) collected from 2014 to 2018. Characteristics of the mother-infant dyads are shown in Table 1. Pregnant women > 18 years old from Swedish-speaking families who received a routine examination at Uppsala University Hospital were invited to participate in the BASIC and BASICchild projects. Only healthy women without a pathological pregnancy were included. Mothers who consented to participate were invited to fill out a series of questionnaires online at 17 and 32 gestational weeks, and postpartum at 6 weeks, 6 months, and 12 months. Mothers and infants who took part in the BASIC Child project visited the Uppsala Child and Baby Lab when the infants were aged 6 (*n* = 118; mean = 185 days, SD = 7.5 days, 59 boys), 10 (*n* = 110; mean = 302 days, SD = 9.2 days, 53 boys), and 18 months (*n* = 104; mean = 544 days, SD = 12.1 days, 53 boys). All infants had a normal 5-minute Apgar score (7-10). The mothers giving birth to the participating infants were 19–41 years old. Sixty-five percent of the mothers held a university degree. Only one mother-infant lived without the second main caregiver.

**Table 1.** Demographic characteristics of participants

| Characteristic | Mother-infant dyad<br>($n = 118$) |
| --- | --- |
| Maternal age, years | 30.54 (3.92) |
| Country of origin |  |
| Scandinavian | 93.1% |
| Other | 6.9% |
| Maternal education |  |
| University or higher | 65.0% |
| Other | 35.0% |
| Cohabiting with the second caregiver | 99.2% |
| With smoking history | 36.4% |
| Employment |  |
| Full-time | 61.2% |
| Part-time | 18.1% |
| Student | 9.5% |
| Sick leave | 4.3% |
| Unemployed | 6.9% |
| Length of gestation, days | 280 (8.09) |
| Infant sex, female | 59% |
| Infant birth weight, g | 3,664 (481) |
| Infant's Apgar score at 5 minutes |  |
| 7 | 0.9% |
| 8 | 2.6% |
| 9 | 6.0% |
| 10 | 90.6% |

*Note: Data are given as the proportion of dyads or mean (SD).*

All procedures in the study were conducted in accordance with the 1964 Declaration of Helsinki ethical standards and approved by the local ethics committee. Mothers who agreed to participate in the online surveys returned their written informed consent prior to the study. For participating infants, all legal guardians provided written consent during each visit prior to the experiment. After each visit to the lab, participants received a gift voucher worth approximately 30 euros.

### Measures of maternal distress

#### Symptoms of depression

The Swedish version of the Edinburgh Postnatal Depression Scale (EPDS) was used to measure symptoms of depression (Cox, Holden, & Sagovsky, 1987; Wickberg & Hwang, 1996). The EPDS includes 10 questions scored from 0 to 3. Thus, the total score ranges from 0 to 30, with higher scores indicating more severe symptoms. The reliability and validity of the EPDS has been shown to be adequate (Affonso, De, Horowitz, & Mayberry, 2000; Cox, Chapman, Murray, & Jones, 1996). Based on validation studies in Swedish samples, the suggestive clinical cut-offs for depressive symptoms are scored more or equal to 13 and 12 during pregnancy and postpartum, respectively (Rubertsson, Börjesson, Berglund, Josefsson, & Sydsjö, 2011; Wickberg & Hwang, 1996). Mothers in the study were invited to complete the EPDS online at 17 and 32 weeks of pregnancy, and postpartum week 6 and month 6.

#### Symptoms of anxiety

Anxiety was measured using the Beck Anxiety Inventory (BAI; Beck, Epstein, Brown, & Steer, 1988). The scale consists of 21 items, with participants indicating the extent to which they were bothered by each item. The score for each item ranges from 0 to 3, with a total score ranging from 0–63. A total score of 0–7 is considered a minimal anxiety level, 8–15 is mild, 16–25 is moderate, and 26–63 is severe (Beck & Steer, 1993). A high level of internal consistency and a good test-retest correlation have been reported (Beck et al., 1988). Mothers in the study completed the online version of the BAI at 17 and 32 weeks of pregnancy and 6 weeks and 6 months of the first postnatal year.

#### Trauma exposure

The Swedish version of the Life Incidence of Traumatic Events was used (LITE; Greenwald & Rubin, 1999; Larsson, 2003). The LITE is a self-reported checklist that consists of 15 fixed items and one optional item. Each item enquires whether the event has occurred, how many times, the age of the first occurrence, and how inconvenient it remains now. The first eight items ask whether different types of non-interpersonal traumatic events (nIP) have occurred, whereas the remaining items ask whether the seven types of events regarding interpersonal traumatic events (IP) occurred. Interpersonal events are defined as events dependent on a conscious act of another human being, such as physical harm, divorce, or separation of parents, etc. Non-interpersonal events include natural disasters, accidents, or illness of others, etc. The sums of occurrences of nIP and IP were used as two variables in the analysis. Acceptable test-retest reliability and validity have been reported (Nilsson, Gustafsson, & Svedin, 2010). As this is an objective measure of occurrences, no clinical cut-off point is reported. Mothers in the current study were invited to complete the LITE online during postpartum 12 months.

### Measure of infants’ attention

Infants’ attention was measured by the look percentage (defined as the total fixation duration of the stimuli divided by the total duration of all tasks at each age) across a variety of free-looking tasks at the age of 6, 10, and 18 months (Table 2). Attention is assumed to be closely linked to oculomotor movement and oculomotor control (Amso & Scerif, 2015; Corbetta et al., 1998) and, in this study, look percentage is used as a proxy for sustained attention (Casey & Richards, 1988; Richards & Turner, 2001). The mean look percentage at 6, 10, and 18 months was 73.63% (SD = 9.84), 73.47% (SD = 9.36), and 79.24% (SD = 6.86), respectively. The Pearson correlation coefficients (Table 3) of attention, look percentage, between different ages were 0.33 (6–10 months, *n* = 110, *p* <0.001), 0.21 (6–18 months, *n* = 103, *p* = 0.04), and 0.31 (10–18 months, *n* = 100, *p* < 0.01), suggesting the stability and internal consistency of attention during the course of development. In the current study, the composite score of look percentage was calculated by averaging each participant’s look percentage measured at three time points and used as the dependent variable. All tasks were recorded using an eye-tracker with a sampling rate of 60 Hz following a 5-point calibration (Tobii TX300, Tobii Technology AB, Sweden).

**Table 2.** Tasks included in look percentage measure from 6, 10, and 18 months of age

| Task description | Test age<br>(months) |
| --- | --- |
| 1. *Give-me gesture interactions* were used to assess action evaluation (Gredebäck & Melinder, 2010; Juvrud et al., 2019). A 40-second context for a give-me gesture followed by appropriate or inappropriate giving was repeated 3 times (26 s total). Four appropriate and four inappropriate trials were presented. | 6, 10 |
| 2. *Change detection task* modified based on Libertus and Brannon (2010) was used to assess the ability to discriminate between numericities. Two image streams were presented to infants simultaneously on both sides of a screen. Images alternated between different numbers of dots with three ratios (1:4, 1:2; 2:3). Each trial lasted for 10 s. | 6, 10 |
| 3. *Multimodal events* were used to evaluate the ability of associative learning (multimodal events bound to locations) (Richardson & Kirkham, 2004). Infants were shown short video clips in which a particular sound was bound to a particular location of a stimulus. | 6, 10 |
| 4. *Biological motion* was used to investigate infants' perception of biological motion (Falck-Ytter et al., 2018). There were two identical animated human-like stimuli presented side-by-side on the screen. One was upright and the other reversed. They showed the same movements but in a reversed mirror direction. There was no auditory stimulation involved. | 6, 10 |
| 5. *Coherent motion task* was inspired by previous studies to measure infants' ability to discriminate between two coherent or random movements (Wattam-Bell, 1994; Wattam-Bell et al., 2010). Two groups of moving dots were presented on two sides of the screen. One contained dots that moved in random directions. | 6, 10 |
| 6. *Gaze following task* was used to examine the degree to which infants follow another person's gaze (Gredebäck et al., 2018; Szufnarowska et al., 2014). | 6, 10, 18 |
| 7. *Pupillary light response* was used to measure the constriction of the pupil diameter in response to a flash of light (Falck-Ytter et al., 2018). | 6, 10 |

Table 2. Continued.
|  | Task description | Test age<br>(months) |
| --- | --- | --- |
| 8. *Small forms discrimination task inspired by previous studies* was used to investigate infants' perception and sensitivity of four geometric forms (Dillon et al., 2013; Izard & Spelke, 2009). In the task, infants were presented with an array of four small forms, each containing two connected lines that formed an angle. Each array included three forms that were identical and one form that deviated. |  | 6, 10 |
| 9. *Face perception* was used to assess whether infants can perceive emotional expressions on faces. Happy, fearful, and neutral facial expressions of three young women were presented to infants at 6 months. An additional two emotions, sad and scared expressions, were presented to infants at 10 and 18 months. All visual stimuli in this task were from the FACES-database (Ebner et al., 2010). |  | 6, 10, 18 |
| 10. *Visual sequence task* was used to examine whether infants can learn the pattern in which the stimuli were presented (Sheese et al., 2008). |  | 10, 18 |
| 11. *Reaching task* was to assess how infants shift their gaze toward a reaching action (Henrichs et al., 2014). |  | 18 |

**Table 3.**
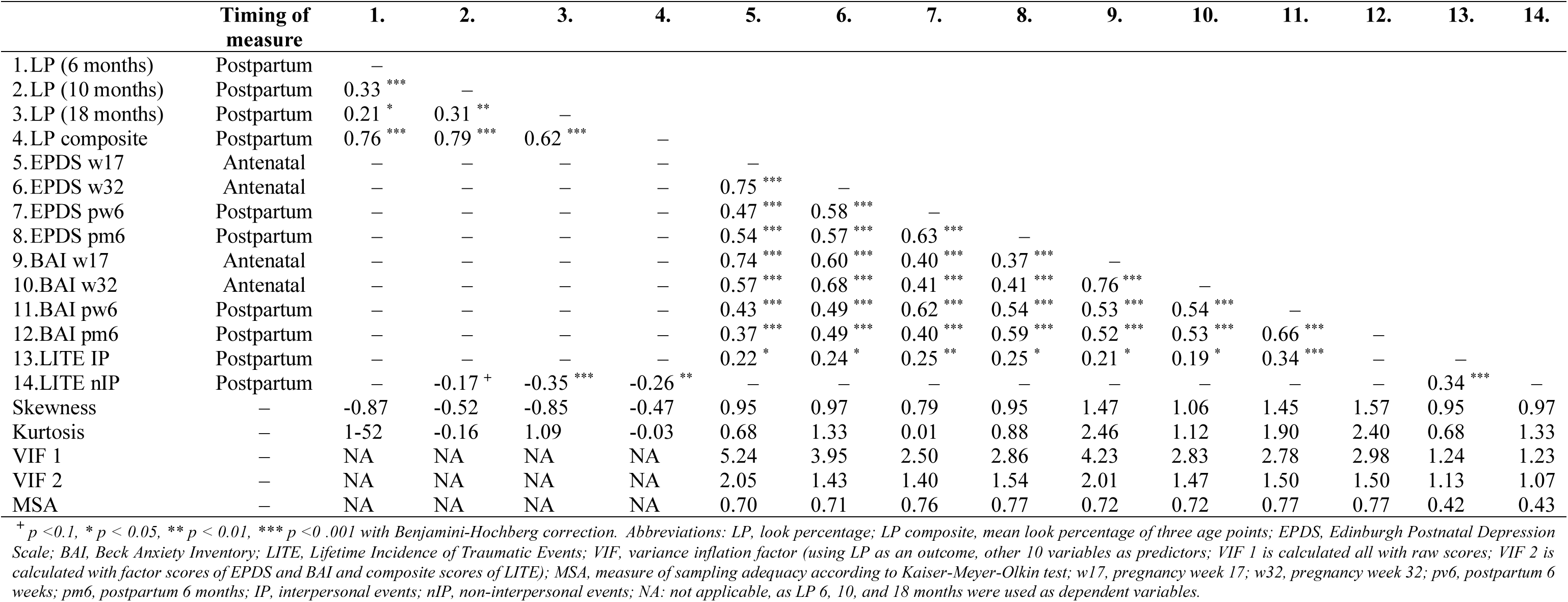
Pearson’s zero order correlations between all variables using raw scores

### Other measurements

In the current study, we preliminarily explore the potential supportive factors suggested by previous studies that may buffer the impact of maternal distress on infants’ attention, whether directly or indirectly. Based on previous literature, several groups of variables measured at antenatal 17 and 32 weeks and postpartum 6 weeks and 6 months were considered. First, subjective delivery expectation and experience (dichotomic rating: positive or negative) was acquired at antenatal 32 weeks and postpartum 6 weeks, respectively. Second, self-reported maternal sleep (measured < 6 hours, 6-8 hours, and > 8 hours) was acquired at all four time points. Third, the interpersonal aspect included interpersonal support and mother-infant bonding. At postpartum 6 weeks and 6 months support included the subjective feeling of partner’s help with a 3-level rating (No; Yes, some help; Yes, much help); subjective feeling of partner sharing the household with a 5-level rating (very little, little, some, much, and pretty much); and partner free from work using a 2-level rating (free from work, not free from work). Bonding was measured based on breastfeeding at postpartum 6 weeks and 6 months and the Postpartum Bonding Questionnaire (Brockington et al., 2001) at postpartum 6 months. Breastfeeding was surveyed using a 3-level rating: Yes, full-time; Yes, also with bottle feeding; No, only with bottle feeding. The Postpartum Bonding Questionnaire was used to detect disorders in the mother-infant relationship. There are 25 questions with total scores ranging from 0 to 125. The higher scores indicate more bonding difficulties perceived by the mother.

### Statistical analysis

We used multivariate linear regression models and a moderator analysis to examine the association between multiple predictors across different time points and the outcome measure. To assess the reliability of the maternal scale instruments, we calculated the internal consistency coefficient, Cronbach’s alpha for each tool: EPDS, 0.87, good; BAI, 0.81, good; and LITE, 0.9, excellent. Before adjusting their scores, the zero-order Pearson correlations (with Benjamini-Hochberg correction), skewness, and kurtosis of all variables were calculated (Table 3). The variance inflation factor (VIF) was calculated based on the assumption that infants’ look percentage is predicted by 10 variables from the EPDS (4 time points), BAI (4 time points), and LITE (1 time point). As seen in Table 3, raw scores for anxiety symptoms during antenatal 17 weeks and postpartum 6 weeks are not in the acceptable range of the kurtosis index. The raw scores of the EPDS, BAI, and LITE did not reach the range of approximate symmetric distribution (kurtosis index acceptable range, -2 to +2; skewness index accetable range -0.5 to + 0.5; Kline, 2015). In addition, the literature has shown that comorbidity of depression and anxiety is common (Dipietro, Costigan, & Sipsma, 2008; Hirschfeld, 2001), so we expected to detect a potential multicollinearity from the raw data. As seen in Table 3, the raw scores of the EPDS and the BAI during antenatal 17 weeks fit the strict criteria for multicollinearity (VIF1 > 4) with other variables (Pan & Jackson, 2008; Rogerson, 2019). Considering non-normal distribution and multicollinearity of the EPDS and BAI, the Kaiser-Meyer-Olkin test was used to examine the sampling adequacy (MSA) and transformed all raw scores from four time points into factor scores (MSA > 0.65; Kaiser, 1974). Missing values were imputed using predictive mean matching (Wulff & Jeppesen, 2017). Individual factor scores of the EPDS and BAI at four time points were calculated using the imputed values. The LITE raw scores, including IP and nIP, were the frequency of the occurrences. They were transformed into dichotomic variables based on the median of the raw scores in order to interpret the interaction. The outcome measure was infants’ look percentage composite. To explore the supportive factors that may act as buffers between maternal distress and infants’ attention, variables listed in the section Other measurements above were analyzed in two directions. One used the infant’s look as the dependent variable. The other used mothers’ distress factor scores from four time points as dependent variables. We used multivariable linear regression models in this exploration.

#### Variable elimination and model fitting

Initially, there was a theoretical selection of 10 predictors included in the current data set that evaluated trauma exposure (1 time point of previous IP and nIP), depressive symptoms (4 time points), and anxiety symptoms (4 time points) in the main analysis to predict infants’ look percentage. No other variables except those listed here have been evaluated as part of the analysis.

In step 1, considering that maternal trauma exposure prior to pregnancy (both IP and nIP) may interact with depression or anxiety, we separated variables into four groups as listed in Table 4 and analyzed four linear regression models independently. Applying a backward stepwise method, the number of variables in each model was reduced (3^rd^ column, Table 4). In step 2, based on the statistical selection shown in Table 4, we combined the significant variables and 2-degree interaction from all models to assess how they jointly predict infants’ attention (measured by look percentage; see Model A, Table 5). Based on Model A, we selected significant variables for Model B (see Table 5). In the third step, we added the sex of infants (Arnett, Pennington, Willcutt, DeFries, & Olson, 2015; Friedman, Bruno, & Vietze, 1974), mothers’ smoking habits (Linnet et al., 2003), education (Clearfield & Jedd, 2013), and the maternal age at birth (Goisis, Schneider, & Myrskylä, 2017) to the analysis (Model C, Table 5).

**Table 4.**
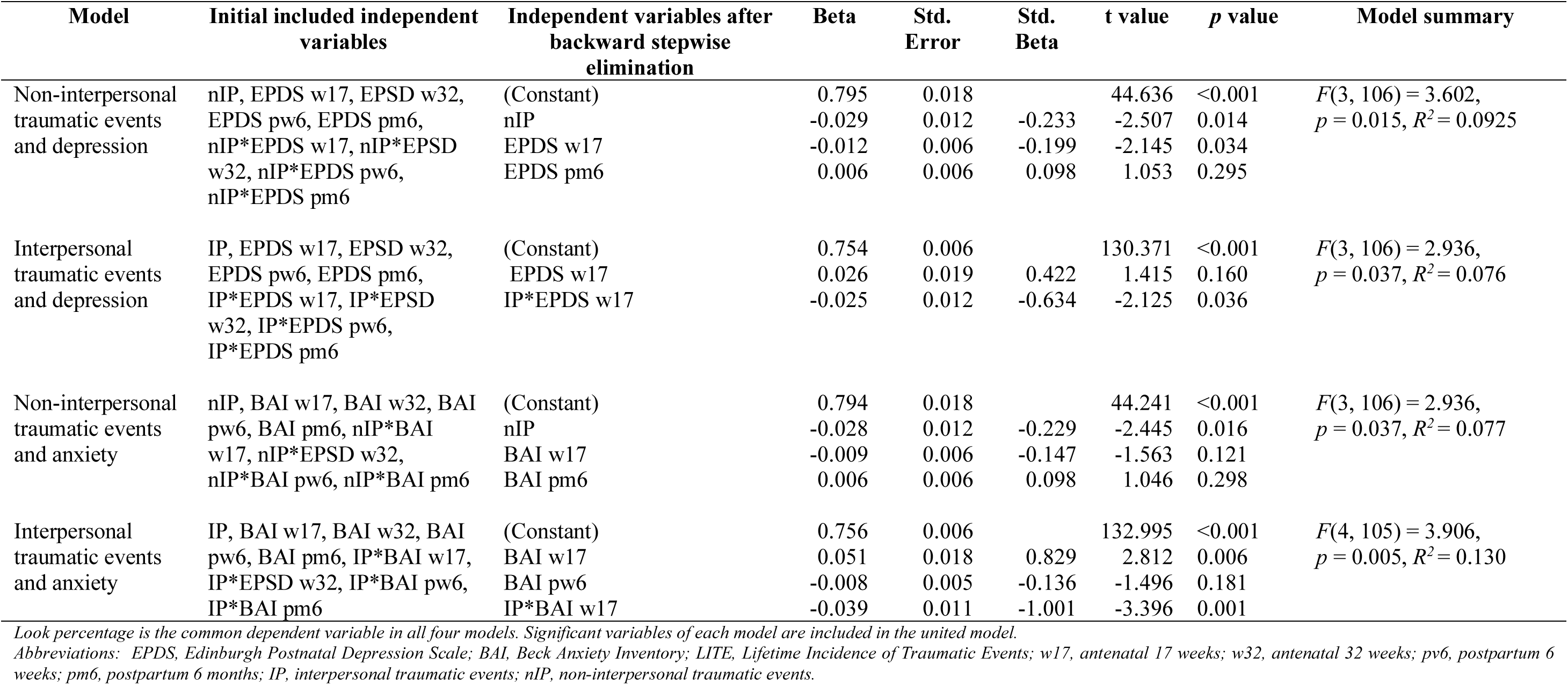
Four separate multivariable linear regression models for systematically selecting variables for the final model

**Table 5.** The final multivariate linear model predicting infants’ look percentage

| Model | Variables | Beta | Std. Error | t value | p value | Model summary |
| --- | --- | --- | --- | --- | --- | --- |
| A | (Constant) | 0.805 | 0.022 | 36.522 | <0.001 | F (6, 103) = 3.698,<br>$p = 0.002$, $R^2$ 0.177 |
|  | nIP | -0.029 | 0.011 | -2.571 | 0.012 |  |
|  | BAI w17 | 0.053 | 0.023 | 2.361 | 0.020 |  |
|  | EPDS w17 | -0.006 | 0.023 | -0.284 | 0.777 |  |
|  | IP | -0.004 | 0.011 | -0.383 | 0.702 |  |
|  | IP*EPDS w17 | -0.001 | 0.015 | -0.079 | 0.937 |  |
|  | IP*BAI w17 | -0.037 | 0.015 | -2.483 | 0.015 |  |
| B | (Constant) | 0.804 | 0.022 | 36.662 | <0.001 | F (4, 105) = 5.287,<br>$p < 0.001$, $R^2$ 0.168 |
|  | nIP | -0.029 | 0.011 | -2.599 | 0.011 |  |
|  | IP | -0.004 | 0.011 | -0.378 | 0.706 |  |
|  | BAI w17 | 0.051 | 0.018 | 2.874 | 0.005 |  |
|  | IP*BAI w17 | -0.039 | 0.011 | -3.534 | <0.001 |  |
| C | (Constant) | 0.820 | 0.055 | 14.791 | <0.001 | F (8, 99) = 2.658,<br>$p = 0.011$, $R^2$ 0.177 |
|  | nIP | -0.029 | 0.012 | -2.495 | 0.014 |  |
|  | IP | -0.002 | 0.012 | -0.152 | 0.880 |  |
|  | BAI w17 | 0.052 | 0.018 | 2.833 | 0.006 |  |
|  | IP*BAI w17 | -0.040 | 0.011 | -3.572 | <0.001 |  |
|  | Infant's sex | 0.001 | 0.012 | 0.50 | 0.961 |  |
|  | Mother's education | 0.022 | 0.014 | 01.588 | 0.115 |  |
|  | Mother's smoking habit | 0.004 | 0.013 | 0.274 | 0.785 |  |
|  | Mother's age at birth | -0.002 | 0.002 | -1.300 | 0.197 |  |

To explore potential supportive factors, additional multivariate linear regressions were performed. In all exploratory analyses, only variables measured before and during the same time point as the dependent variable were included. The backward stepwise variable elimination method was applied to select variables.

All tests were two-sided tests with *p* < 0.05 considered significant. All statistical analyses were performed using R 4.0.3 (Team, 2020).

## Results

### Infants’ attention predicted by maternal anxiety symptoms and childhood trauma

#### Multivariate regression analyses

As seen in Table 5, Model A (*F*(6, 103) = 3.698, *R*^*2*^ = 0.177, *p* = 0.02) includes all significant variables systematically selected from Table 4 as described in the Methods. We observed that higher levels of interpersonal traumatic experience in childhood interact with anxiety during the 2^nd^ trimester and a decrease in infants’ attention (see Model A in Table 5, *b* = - 0.037, *p* = 0.015). We also found two main effects. First, when mothers were exposed to higher levels of non-interpersonal trauma in childhood, there was a decrease in infants’ attention to audio-visual stimuli (*b* = - 0.029, *p* = 0.012). Second, when mothers reported higher levels of anxiety during the 2^nd^ trimester, infants increased their attention (*b* = 0.053, *p* = 0.02).

The second step, Model B (*F*(4, 105) = 5.287, *R*^*2*^ = 0.168, *p* < 0.001) contained only variables that were significant predictors in Model A. All effects remained significant in Model B: the interaction between interpersonal traumatic events and anxiety level during the 2^nd^ trimester (*b* = - 0.039, *p* < 0.001), the main effect of non-interpersonal traumatic events (*b* = - 0.029, *p* < 0.05), and the main effect of anxiety level during the 2^nd^ trimester (*b* = 0.051, *p* < 0.01).

After controlling for infant’s sex, mother’s education, smoking habits, and maternal age at birth, Model C (*F*(8, 99) = 2.658, *R*^*2*^ = 0.177, *p* = 0.011) showed that the interaction between interpersonal traumatic experiences and anxiety during the 2^nd^ trimester (*b* = - 0.040, *p* < 0.001), the main effect of non-interpersonal traumatic events (*b* = - 0.029, *p* < 0.05), and the anxiety level during pregnancy during 2^nd^ trimester (*b* = 0.052, *p* < 0.01) all remained significant. Figure 1 visualizes the results of Model C.

**Figure 1.**
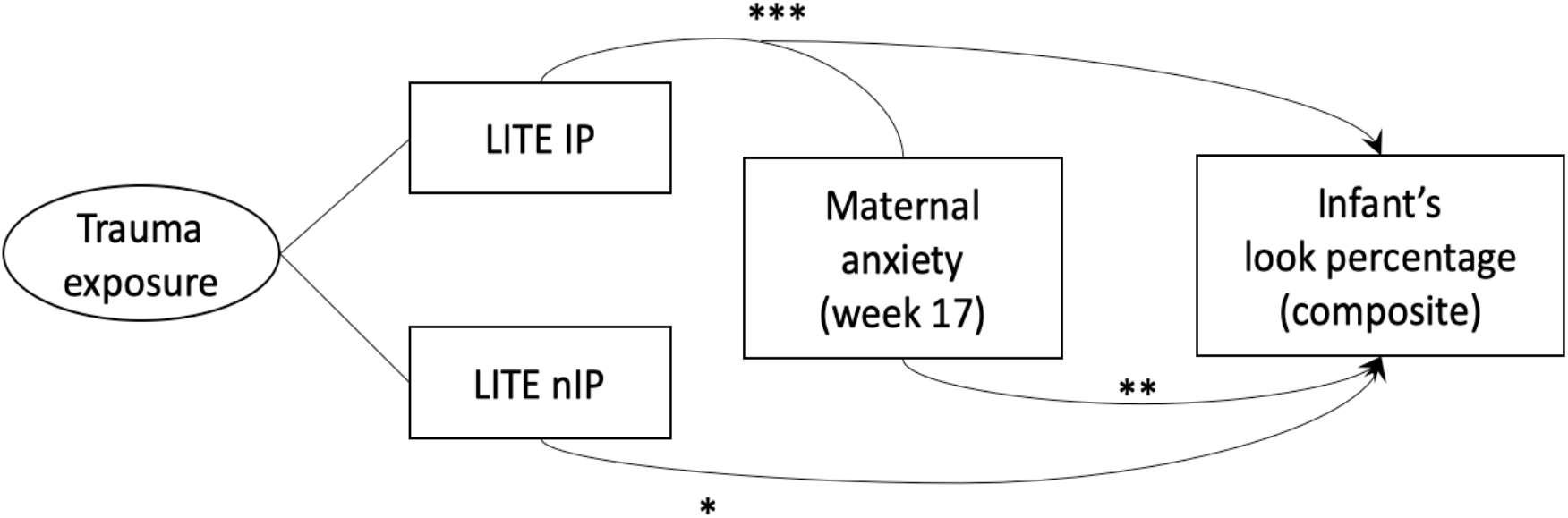
Illustration of the multivariate linear regression model after adjusting for the sex of the infant, mother’s education level, smoking history, and the maternal age at birth. Non-interpersonal traumatic experiences in mother’s childhood and maternal anxiety in early pregnancy had a direct impact on the infants’ look percentage (p = 0.014 and 0.006, respectively). When anxiety at week 17 of pregnancy interacts with interpersonal traumatic exposure in childhood, the negative impact on the infants’ look percentage is highly significant (p <0.001). LITE, Lifetime Incidence of Traumatic Events; IP, interpersonal events; nIP, non-interpersonal events.

#### Moderation analysis

Following the results described above, exposure to interpersonal traumatic events in childhood was examined as a moderator of the relationship between the anxiety level during the 2^nd^ trimester and the infants’ look percentage after adjusting for infant sex and mother’s education. Figure 2 displays the slopes for the anxiety level during antenatal 17 weeks and the levels of the exposure to interpersonal traumatic events predicting infants’ attention. As indicated by the change in the direction, the effect is moderated by interpersonal traumatic events (F(5, 103) = 2.916, *R*^*2*^ = 0.124, *p* = 0.018). In other words, with a higher exposure of interpersonal traumatic events, infants’ attention decreases as the mother’s anxiety level increases.

**Figure 2.**
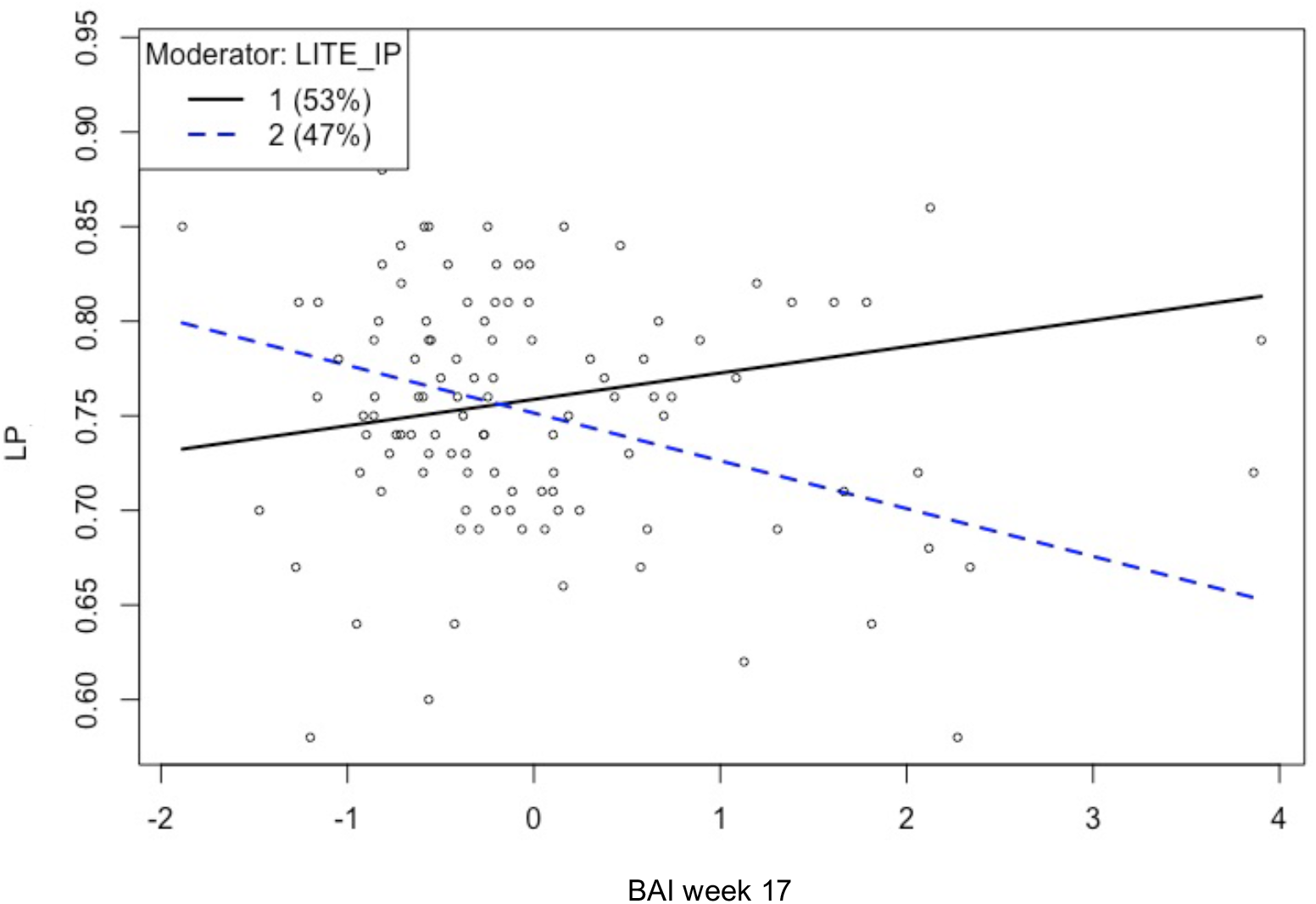
The relationship between maternal anxiety (Beck Anxiety Inventory at antenatal 17 weeks, BAI week 17) and infants’ attention, look percentage (LP), is moderated by the level of interpersonal traumatic events (IP) in childhood measured by Life Incidence of Traumatic Events (LITE). Level l (solid line) represents mothers who exposed to less trauma in childhood compared to those at level 2 (dotted line).

### Exploratory findings of supportive factors against the negative impact of maternal distress on infants’ attention

To explore the supportive factors that may act as buffers between maternal distress and infants’ attention, variables regarding three aspects were analyzed. As seen in Table 6, we retained two variables from three aspects (i.e., subjective delivery expectation and experience, maternal sleep, and interpersonal aspect) based on the backward stepwise method. We did not observe any significant direct impact of these factors on infants’ attention (*F*(2, 103) = 2.729, *R*^*2*^ = 0.05, *p* = 0.069). However, when controlling for the interpersonal traumatic experience, we observed that different supportive factors significantly ease mothers’ anxiety levels at different time points. In Table 7, we summarize each anxiety measure time point with one model that includes protective variables selected by the backward stepwise method. During the 2^nd^ trimester, there is a tendency for positive expectation for delivery to be associated with lower anxiety levels (*b* = -0.766, *p* = 0.003), though the model is not significant (*F*(5, 101) = 2.077, *R*^*2*^ = 0.09, *p* = 0.07). During the 3^rd^ trimester, the positive expectations for delivery (*b* = -0.230, *p* = 0.034) and more sleep hours (*b* = -0.603, *p* = 0.034) measured in the same period significantly reduced anxiety (*F*(6, 100) = 3.08, *R*^*2*^ = 0.16, *p* = 0.008). In the same model (during the 3^rd^ trimester), more sleep hours during the 2^nd^ trimester increased the anxiety level during the 3^rd^ trimester (*b* = 0.341, *p* = 0.036). Later, during postpartum 6 weeks, a positive delivery experience significantly eased the anxiety level (*b* = -0.770, *p* = 0.033). Breastfeeding had significant positive effects on reducing maternal anxiety at both postpartum 6 weeks (*b* = -0.319, *p* = 0.036) and 6 months (*b* = -0.328, *p* = 0.025), though the latter model (*F*(7, 92) = 1.858, *R*^*2*^ = 0.12, *p* = 0.086) only approached significance (see Table 3).

**Table 6.** Exploratory results of multivariable linear regression predicting look percentage using potential protective factors

| Variables | Estimate | Std.<br>Error | t value | p value | Model summary |
| --- | --- | --- | --- | --- | --- |
| (Constant) | 0.733 | 0.013 | 56.101 | < 0.001 | $F(2, 103) = 2.729$;<br>$p = 0.069$, $R^2 = 0.05$ |
| Partner free from work measured during postpartum 6 months | 0.028 | 0.017 | 1.548 | 0.125 |  |
| Sleep assessed at gestational 32 weeks | 0.011 | 0.007 | 1.6 | 0.113 |  |

**Table 7.** Exploratory results of multivariable linear regression buffering the impact of anxiety across four time points using supportive factors

| Anxiety time point | Variables | Estimate | Std. Error | t value | p value | Model summary |
| --- | --- | --- | --- | --- | --- | --- |
| Antenatal 17 weeks | (Constant) | -0.103 | 0.625 | -0.166 | 0.869 | $F(5, 101) = 2.077$;<br>$p = 0.07$, $R^2 = 0.09$ |
|  | Positive expectation for delivery | -0.603 | 0.287 | -1.669 | 0.034 |  |
|  | Sleep 17w | 0.204 | 0.167 | 1.219 | 0.226 |  |
|  | Interpersonal traumatic experience | 0.220 | 0.198 | 1.110 | 0.270 |  |
| Antenatal 32 weeks | (Constant) | 0.531 | 0.566 | 0.938 | 0.350 | $F(6, 100) = 3.08$;<br>$p = 0.008$, $R^2 = 0.16$ |
|  | Positive expectation for delivery | -0.766 | 0.255 | -2.999 | 0.003 |  |
|  | Sleep 17w | 0.341 | 0.160 | 2.128 | 0.036 |  |
|  | Sleep 32w | -0.230 | 0.108 | -2.129 | 0.036 |  |
|  | Interpersonal traumatic experience | -0.071 | 0.178 | -0.401 | 0.689 |  |
| Postpartum 6 weeks | (Constant) | 2.312 | 0.839 | 2.757 | 0.007 | $F(9, 85) = 2.888$;<br>$p = 0.005$, $R^2 = 0.23$ |
|  | Positive experience for delivery | -0.770 | 0.356 | -2.165 | 0.033 |  |
|  | Partner free from work pw6 | 0.428 | 0.266 | 1.610 | 0.111 |  |
|  | Partner assists household pw6 | -0.103 | 0.093 | -1.104 | 0.273 |  |
|  | Sleep 32w | 0.089 | 0.121 | 0.732 | 0.466 |  |
|  | Sleep pw6 | -0.106 | 0.076 | -1.400 | 0.165 |  |
|  | Breast feeding pw6 | -0.319 | 0.150 | -2.129 | 0.036 |  |
|  | Interpersonal traumatic experience | 0.017 | 0.220 | 0.070 | 0.945 |  |
| Postpartum 6 months | (Constant) | 1.254 | 0.711 | 1.764 | 0.081 | $F(7, 92) = 1.858$;<br>$p = 0.086$, $R^2 = 0.12$ |
|  | Partner free from work pw6 | 0.338 | 0.277 | 1.219 | 0.226 |  |
|  | Partner general support pw6 | -0.211 | 0.163 | -1.293 | 0.199 |  |
|  | Sleep 17w | -0.283 | 0.166 | -1.710 | 0.090 |  |
|  | Breast feeding pw6 | -0.328 | 0.144 | -2.283 | 0.025 |  |
|  | Interpersonal traumatic experience | 0.124 | 0.201 | 0.617 | 0.538 |  |

Note: 17w, antenatal 17 weeks; 32w, antenatal 32 weeks; pw6, postpartum 6 weeks; pm6, postpartum 6 months. All models control for infant sex and mother's education and interpersonal traumatic exposure in childhood.

## Discussion

Attention in infancy is fundamental to cognitive development and learning (Posner & Rothbart, 2009) and is linked to language development (Yu et al., 2019), social behavior (Bowers et al., 2019), and academic skills in childhood (Shannon et al., 2021). The primary goal of the current study was to investigate whether maternal distress affects the development of attention in infancy. We found that exposure to non-interpersonal and interpersonal traumatic experiences childhood has cross-generational effects on infants’ attention. Moreover, childhood interpersonal trauma experience moderates the maternal anxiety level during the 2^nd^ trimester and hinders the development of attention in infancy. In other words, our results indicate that both traumatic experiences before pregnancy and perinatal anxiety increase the vulnerability of mothers and elevate the risk for poor attention of infants. From the standpoint of prevention, our findings underscore the importance of early screening and intervention for mental health issues to support mothers and infants and prevent long-term consequences, even before the pregnancy starts.

In our exploration of supportive factors, we did not observe any of these factors as being related to the development of attention. Our small sample size may limit the observation of this effect, or the effects may only be measurable at a later time point. In addition, the effects may be related to other supportive factors we did not include. Though we observed no supportive factor associated with infants’ attention, our results demonstrate that maternal anxiety itself is diminished by several supportive factors. For example, positive delivery expectations and experience, strong mother-infant bonding, and sufficient sleep may contribute to the mother’s well-being and indirectly support the infant’s development of attention. Although more systematic and longitudinal studies are needed to further link the supportive factors and infants’ attention, our preliminary findings help pinpoint the possibility of facilitating the development of attention in infancy through early support of mothers’ mental health.

In our longitudinal data across pregnancy to early infancy, we found a particular vulnerability in the 2^nd^ trimester. There are two main possible explanations for this finding. The first explanation is that the fetal brain is vulnerable to the in-utero environment due to the critical period of neurogenesis. Exposure to risk factors during this stage of development leads to altered neuron connectivity. Compared to the 1^st^ and 3^rd^ trimesters, exposure to ethanol in the 2^nd^ trimester has been reported to cause great neuronal loss in rodents (Miller & Potempa, 1990), attenuated cerebral blood flow (Mayock, Ness, Mondares, & Gleason, 2007), and long-lasting alternations in synaptic plasticity (Helfer, White, & Christie, 2012) in the human fetus. In children, Buss et al. (2010) also reported that maternal anxiety during the 2^nd^ trimester, but not later during pregnancy, is associated with gray matter reduction in several brain areas in children (6–9 years old), including the prefrontal lobe, which is a crucial area in cognitive development (B. Casey, Tottenham, Liston, & Durston, 2005) and controls attention (Paneri & Gregoriou, 2017). The second explanation is associated with the elevated cortisol levels in mothers during pregnancy. Compared to mothers who do not experience maternal anxiety, childhood maltreatment, or an adverse environment, mothers who experience anxiety or these traumatic experiences have a higher level of cortisol (Bowers et al., 2018; Leff-Gelman et al., 2020; Stephens et al., 2021). Scheinost et al. (2020) reported that increased cortisol levels during the 2^nd^ trimester and increased subjective maternal distress in the 3^rd^ trimester are associated with weaker connectivity of the anterior cingulate cortex of neonates (Scheinost et al., 2020). The anterior cingulate cortex has been linked to infant’s attention (Reynolds, Courage, & Richards, 2010; Reynolds & Richards, 2005) and ADHD in children (Kelly et al., 2009; Qiu et al., 2011) and adults (Seidman et al., 2006). In addition, one previous study investigating infants’ cognitive development at 12 months of age reported that infants with higher cognitive performance were born to mothers with lower maternal cortisol levels in the 2^nd^ trimester but higher cortisol levels in the 3^rd^ trimester (Davis & Sandman, 2010).

Interestingly, maternal depression showed no association with infants’ attention. However, previous literature has shown that infants of depressed mothers have a less synchronous gaze in the mother-infant interaction (Lotzin et al., 2015; Væver, Krogh, Smith-Nielsen, Christensen, & Tharner, 2015) that may affect the development of attention (Niedźwiecka et al., 2018). Similar to two well-controlled studies investigating cognitive development, maternal depression during pregnancy and infancy did not affect cognitive development at the age of 3 years (Tse, Rich-Edwards, Rifas-Shiman, Gillman, & Oken, 2010) and 18 months (Piteo, Yelland, & Makrides, 2012), respectively. In the context of the current study, there are several plausible reasons for this finding. First, the association between maternal depression and infants’ attention may not exist. However, using the same dataset investigating gaze following, infants of mothers with lower levels of postpartum depression presented better skills in synchronizing visual attention with others based on their gaze direction (Astor et al., 2020). Though mutual gaze interaction can predict attention in infancy (Niedźwiecka et al., 2018), our data and Astor et al.’s (2020) study show that there may be more than one pathway of mother-infant interaction that influences the development of attention. Second, it is possible that the impact of maternal depression on infants’ attention is cumulative and becomes significant only in childhood (Wang & Dix, 2017). Third, as maternal depression is complex and heterogeneous in nature (Mughal et al., 2018; Santos, Tan, & Salomon, 2017; Wikman et al., 2020), our four time points may not reflect the complexity and heterogeneity of associations across mothers and infants. Lastly, because of the rigorous nature of the BASIC study, among mothers with depressive symptoms, a higher proportion of those with high functioning/cognitive skills (of which the children might also have good attention) are hypothesized to have filled out the questionnaires, introducing a possible selection bias.

Keeping these alternatives in mind, we cautiously propose another reason. Given the high comorbidity of depression and anxiety in our data (Table 3) and the literature (Dipietro et al., 2008; Hirschfeld, 2001), we propose that anxiety may be the driving force behind peripartum depression. For example, when we examined depressive and anxiety symptoms separately (Table 7), they showed a unique effect during the 2^nd^ trimester. When we further combined all dimensions and examined the effect while simultaneously controlling others, anxiety dominated the effect. To the best of our knowledge, maternal depression and anxiety are rarely combined and related to child development, meaning that the importance of maternal anxiety may have been interpreted as an effect of depression in prior work. However, the complexity and dynamics between traumatic experiences, depression, and anxiety and how the dynamics change over time are beyond the scope of the current study. Future studies are needed to help us understand how maternal mental health affects infants’ attention. Most importantly, it will provide us more knowledge on promoting maternal mental health and infant development.

Finally, and especially due to our limited sample size, our results must be interpreted in light of some limitations. Firstly, we could not compare clinically severe cases due to the relatively small number of severely depressed mothers. To deal with the relatively small sample size and the significant collinearity between depression and anxiety, we calculated the factor scores for depression and anxiety separately at four different time points. This may prevent the plausible interactions at different stages and different levels from being observable in our results. Moreover, our sample is limited to a homogenous population in Uppsala (Sweden), with more than half of participating mothers having education levels of university or higher. In addition, we did not control for the possible influence of partners’ mental health on mothers’ well-being and infants’ attention. As our results indicate the important influence of interpersonal traumatic experiences, future studies should consider this interpersonal aspect and its dynamics with regard to mothers’ well-being.

A strength of this study is that it uses a multi-dimensional approach and investigates the distinct impact of maternal distress during pregnancy and postpartum. Second, the robust attention measure provides a reliable and steady observation of individual differences in the development of attention.

Our findings add to the growing body of research, suggesting that prevention and intervention should start before pregnancy for both mothers and infants. Lastly, the findings describe a previously undocumented connection between maternal early trauma, anxiety, and the development of attention in infants. Treating pregnant women’s anxiety, especially if she has experienced traumatic events in the past, may not only improve the lives of mothers, but also support positive development of their children from infancy onwards.

## Data Availability

All relevant information is available on the link provided below.

https://nyu.databrary.org/volume/828

